# A systematic review of the effectiveness and implementation readiness of psychosocial interventions for psychosis in South Asia

**DOI:** 10.1101/2023.03.09.23287063

**Authors:** Sophie Lyles, Zahra Khan, Onaiza Qureshi, Madiha Shaikh

## Abstract

**Background:** Little is known about the effectiveness and implementation of psychosocial interventions for psychosis in low- and middle-income countries (LMICs). In South Asia, specialist psychiatric resources are scarce. Support for psychosis often falls on the family or caregiver which can increase feelings of burden, impact caregivers’ wellbeing, and increase mental health stigma. Psychosocial interventions are increasingly used for psychosis in South Asia and could reduce relapse and symptoms, reduce caregiver burden, conserve cost and resources. The aim of this review was to appraise the effectiveness and implementation readiness of psychosocial interventions for people with psychosis in LMICs within South Asia.

**Method:** A systematic search was conducted on MEDLINE, PsycINFO, Global Health, and Web of Science. The review was prospectively registered on PROSPERO (CRD42022329254). Studies were rated on two scales assessing quality and implementation readiness.

**Results:** Twenty-six papers were included, nine intervention-types including community-based interventions/assertive outreach; CaCBTp; FAP; psychoeducation; cognitive retraining/rehabilitation; social cognition/skills; family/ caregiver intervention; telehealth intervention; yoga-based intervention in six South Asian countries.

Findings suggest a multicomponent community-based intervention (MCBI) was the most implementation ready due to its standardisation, good clinical outcome outcomes for patients and caregivers, and training and cost evaluations.

**Conclusion:** Of the included studies, MCBI and community-based outreach interventions utilising lay health workers appear to be the most implementation ready and are suggested to best address the treatment gap in South Asia.

## 1. Introduction

Around 21 million people live with schizophrenia, with the majority residing in low and middle-income countries (LMICs) (1). The rates of mental health disorders in South Asia are typically under-reported due to data scarcity, and a significantly more limited focus on the treatment of mental health disorders (2). Mental health disorders typically place a significant psychological, social and economic burden on South Asian citizens, however, up to 90% of people with mental illnesses who need treatment, do not receive it (3). Inadequate infrastructure, lack of resources, trained specialist staff or cultural adaptations, focus on biomedical approaches, potential of transferability for effective and scalable interventions given similar cultural determinants and healthcare structures shed light on the significant treatment gap in LMIC and within South Asia (4, 5). Treatment outcomes, particular global functioning and negative symptoms can be enhanced by psychosocial interventions aimed at social, health and economic needs of the individual (6). Disease Control Priorities (DCP-3) suggest self-help, support groups, community-based rehabilitation and family therapy or support that could address discrepancies in the implementation of such interventions in LMICs (7). A task shifting model can address the lack of specialist mental health workers by utilising non-specialist workers such as lay health workers, peer support workers and nursing staff without psychiatric training. Given psychosocial interventions have a significant effect on hospitalizations, functioning and relapse rates (8), and aim to address broader social and life-skills (9), they are an integral part of recovery to enable individuals and caregivers to reach their goals.

Research suggests there is notably more evidence for the feasibility and acceptability of psychosocial interventions for psychosis in South Asia than for the effectiveness of such interventions (10).

The aim of this review is to assess effectiveness and implementation readiness (IR) of psychosocial interventions for people with psychosis on patient outcomes in South Asia.

## 2. Method

### 2.1 Design

A systematic search of published research on psychosocial interventions for people with psychosis in South Asia was conducted. The review was prospectively registered on PROSPERO (CRD42022329254). The PRISMA systematic procedure was followed to conduct the search, screening, and synthesis of findings (11) (Figure 1).

**Figure 1.**
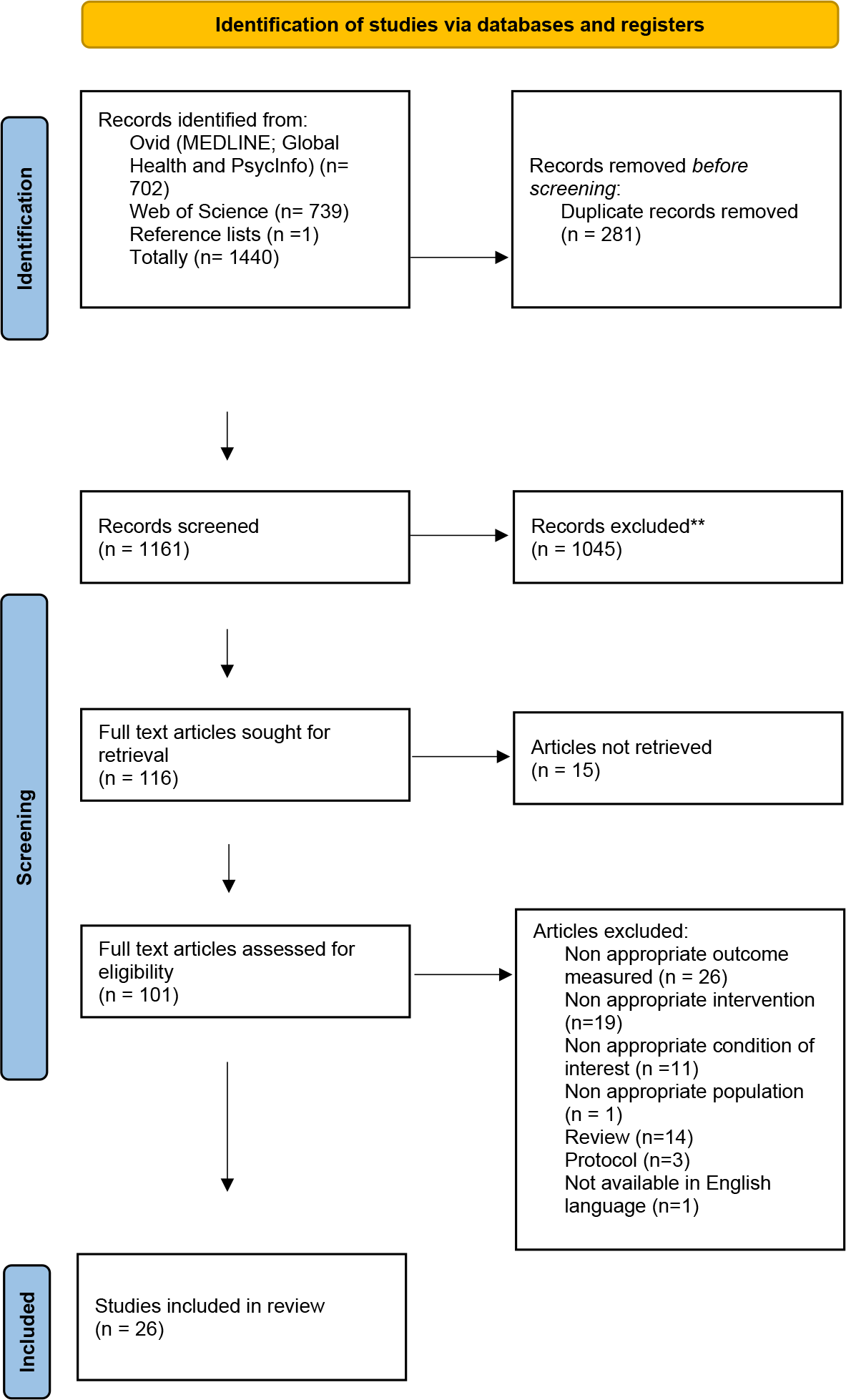
PRISMA flow diagram.

### 2.2 Search strategy

Reviewers searched MEDLINE, PsycINFO, Global Health, Web of Science, review articles and reference lists of eligible studies. The search was conducted on 15 October 2022 and all years were included. Search terms for psychosis included ‘schizoaffective’ or ‘schizo*’ or ‘psychosis’ or ‘psychoses’ or ‘psychotic’. Intervention search terms were: ‘Psychosoc*’ or ‘Family intervention*’ or ‘CBTp’ or ‘Social skills training’ or ‘Psychological therap*’ or ‘Social therap*’ or ‘Non pharma*’ or ‘Caregiver intervention*’ or ‘intervention*’ or ‘psycho therap*’ or ‘cognitive therap*’ or ‘cognitive behavioural therap*’ or ‘counselling*’ or ‘psychotherap*’. Search terms for South Asia were: ‘South Asia*’ or ‘India*’ or ‘Bangladesh*’ or ‘Bhutan*’ or ‘Maldiv*’ or ‘Nepal*’ or ‘Pakistan*’ or ‘Sri Lanka*’ or ‘Afghan*’ or ‘Indoasia*’ or ‘Indo asia’.

### 2.3 Eligibility Criteria

Any psychosocial intervention with a focus on psychological and/or social factors as opposed to biological factors (pharmacological interventions) delivered to people with psychosis or their caregivers with the aim of improving individual outcomes in South Asian countries: Afghanistan, Pakistan, India, Bhutan, Maldives, Nepal, Sri Lanka, Bangladesh was included. Papers were excluded if no full text was available in English.

### 2.4 Screening and Selection

Papers were imported to Mendeley, where duplicates were removed, and titles and abstracts were screened against the eligibility criteria. Full texts were retrieved and screened against the eligibility. References lists were sought for any further studies meeting the inclusion criteria.

### 2.5 Data Extraction

Data reporting on country, design, number of participants, duration of follow up (FU) and intervention characteristics such as content and outcomes were extracted.

### 2.6 Quality Assessment and Implementation readiness check

Implementation readiness was assessed via the ImpRess (12) checklist. This is a 26-item checklist assessing motivation, theory of change, implementation context, experience, employee and manager support and resources. Downs and Black (13) checklist was used to assess quality of papers. This consists of 27-items to assess the quality of randomised and non-randomised research. All studies were rated by at least 2 independent raters (SL, MS, ZQ, OQ) on both checklists. Discrepancies were resolved through discussion or by a third rater.

### 2.7 Data analysis and synthesis

Due to heterogeneity (of populations and interventions examined, research methods employed, and outcomes measured) of included studies fa sound meta-analysis could not be conducted. Therefore, to ensure that the results could be reported in a systematic manner, a narrative synthesis was conducted. The narrative synthesis included: the intervention modality, outcomes targeted and the nature and duration of the intervention’s delivery along with the results of each intervention’s effectiveness. A summary is presented in Table 1.

**Table 1.**
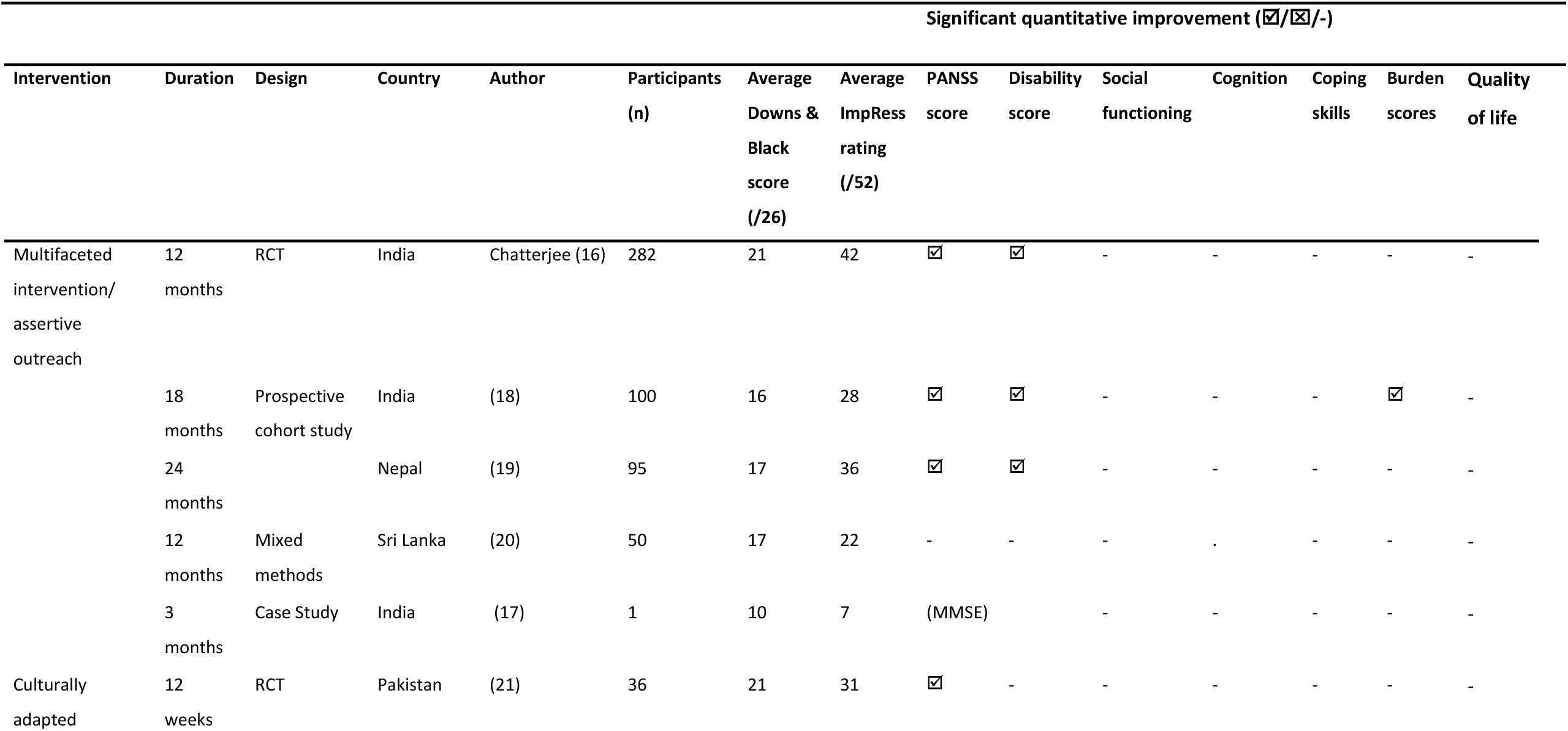
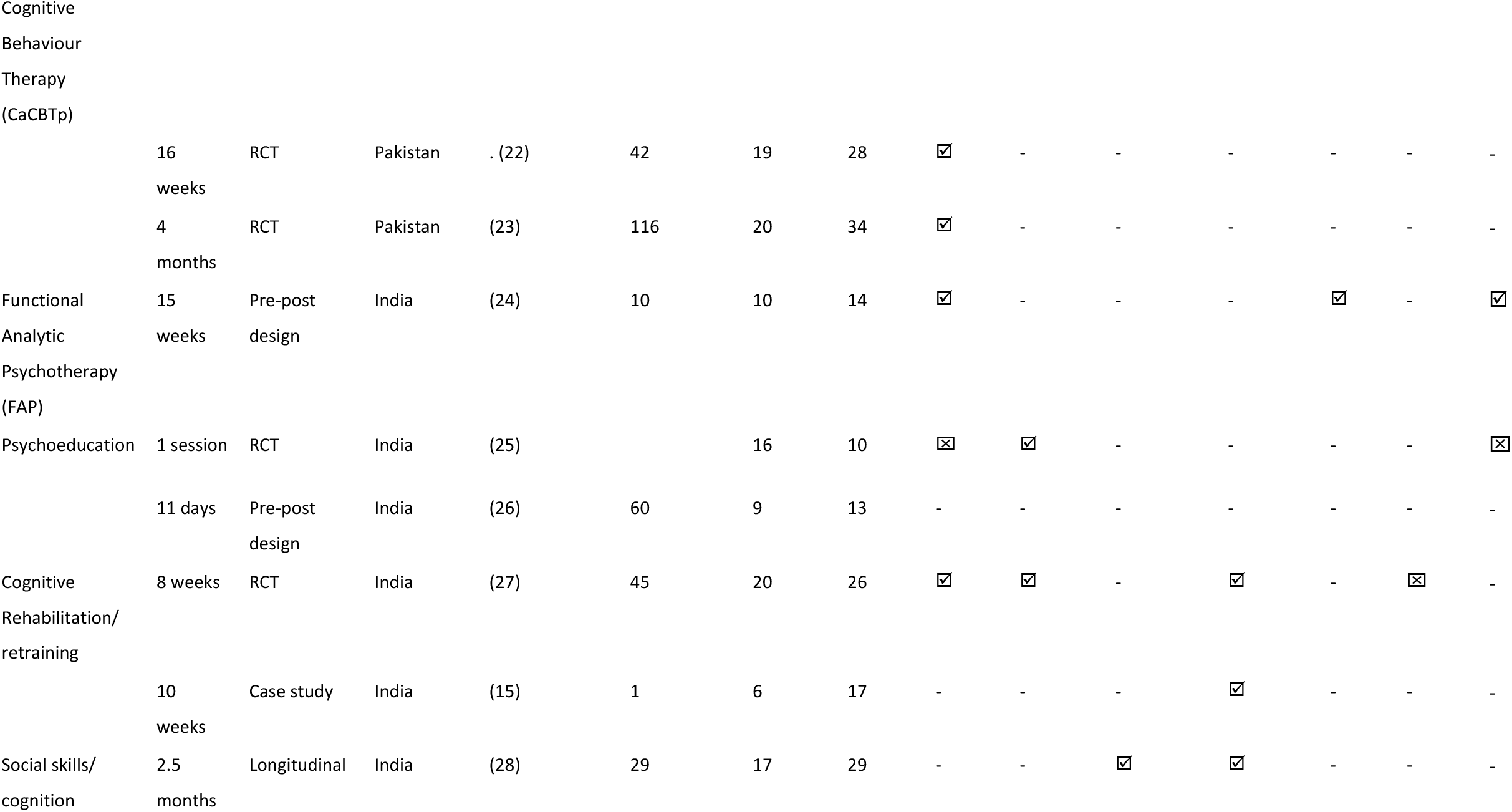
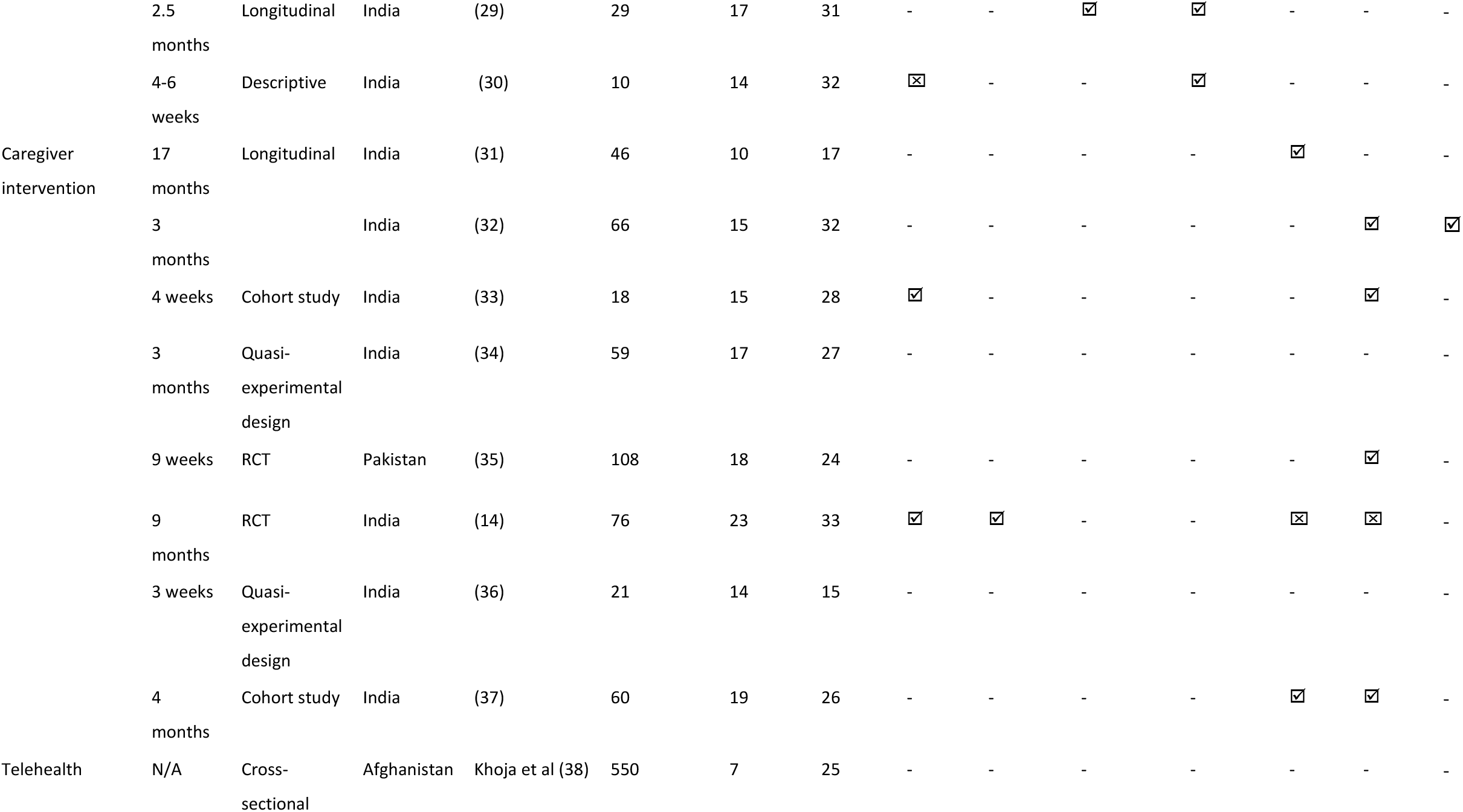
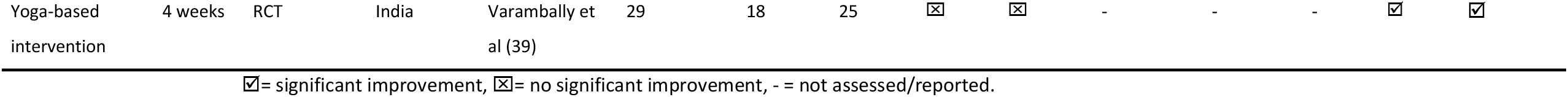
Summary of included studies

## 3. Results

A total of 1161 records were screened following the removal of duplicates. Title and abstract screening excluded a further 1045, leaving 116 left for retrieval. No full text was available for 15 papers and 75 studies were excluded for reasons outlined in figure 1. This left 26 studies to be included in this review encompassing 9 interventions across 6 countries.

### 3.1 Quality Assessment

Downs and Blacks (13) ratings averaged at 16/26. The highest quality study was an evaluation study of a psychoeducation intervention by Kulhara et al (2009) (14) with 23/26 and the lowest quality study was Rajeswaran, Taksal and Jain (15) with 6/26.

Agreement between raters was substantial (ƙ= .690). ImpRess (IR) ratings averaged at 26/52. A multicentre, parallel-group, randomised controlled trial in India scored highest (42/52) (16) and the least was a cognitive rehabilitation programme in India (17), scoring 7/52. Studies scored highest in motivation for evaluating the intervention, the theoretical underpinnings of how and why change could occur and population characteristics. Few studies reported on managerial and employee support, training costs, time needed for planning and setting up the intervention or barriers to implementation. Agreement between raters was substantial (ƙ = .781).

Studies were grouped together based on type of intervention (community-based interventions/assertive outreach; CaCBTp; FAP; psychoeducation; cognitive retraining/rehabilitation; social cognition/skills; family/ caregiver intervention; telehealth intervention; yoga-based intervention).

### 3.2 Comprehensive community-based interventions/ Assertive Outreach

#### 3.2.1 India

A randomised-controlled trial (RCT) compared the effectiveness of a community-based intervention and facility-based intervention (n=187), to facility-based care alone (n=95) (16). It was awarded the highest IR score (42/52) due to the consideration of cost-effectiveness, training community health workers and tailoring the intervention to the individual needs of the person. It comprised of structured needs assessments; psychoeducation for participants and caregivers; adherence support; health promotion support; individualised rehabilitation to address personal, social, and work functioning; support with experiences of stigma and discrimination; self-help and user-led support; access to wider network to address contextual social issues such as benefits, inclusion, and employment. Participants with a primary diagnosis of schizophrenia were recruited from three rural sites. After 12 months, Positive and negative symptom scale (PANSS) ((40) (p= 0.08) and Indian disability evaluation and assessment scale (IDEAS) (41) scores (p= 0.01) were lower in the experimental group (16).

Murthy et al., (18) delivered a community outreach intervention comprised of memory, attention, planning, verbal and non-verbal fluency and visuo-spatial tasks for 8 weeks. Treatment as usual (TAU) comprised of usual drug treatment and psychoeducation. Retraining significantly improved cognition across all domains and WHO disability assessment scale (p<.001). PANSS Negative scores were significantly reduced (p= 0.019)

Sarwar & Bashir (17) conducted a case study of an individual with chronic schizophrenia residing in a rehabilitation unit. Symptoms were evaluated using mini mental state examination, however this lacked standardisation and applicability to other contexts. The intervention consisted of a 3-month management plan aiming to increase quality of life and socialization. They measured each problem: hygiene, socialization, and odd patterns of behaviour on a 0– 10 Likert-scale (10= problematic; 0= not problematic). They found a reduction in ratings for all domains post-intervention.

#### 3.2.2 Nepal

As part of the Programme for Improving Mental Health Care (PRIME), a primary-care-based mental health service receiving a package of care including adherence support and psychological treatment by primary care mental health workers was evaluated. At 12-months, there was a significant reduction in PANSS scores (p<.001) which was sustained at FU after 24-months (p<.001). WHODAS scores significantly improved (p<.0.05) which continued to improve at FU (p<.001) (19).

#### 3.2.3 Sri Lanka

A study by Rodrigo et al (20) evaluated rapport building interviews, follow-up phone calls, home visits and ongoing needs assessments conducted by a mental health development officer (MHDO). MHDOs are recruited from non-psychiatric university courses to mimic the role of a psychiatric social worker. Despite the outcome indicators not differing at a level of statistical significance, the intervention group had better clinic attendance, less relapses, and less defaults.

### 3.3 Culturally adapted Cognitive Behaviour Therapy for psychosis (CaCBTp)

#### 3.3.1 Pakistan

A RCT evaluated a 12-week CaCBTp intervention against TAU (21). They followed the manual developed by Kingdon and Turkington (42) encompassing collaborative understanding, exploring antecedents, normalising rationale, treating co-morbid anxiety and depression and case formulation as well as specific techniques to address positive symptoms to explore and develop coping strategies. The cultural adaptations consisted of using culturally appropriate idioms to explain concepts relating to symptoms and causes. For example, referring to religious teachings, speaking in native language (Urdu) and involving carers and family. At 3-months, the CaCBTp group’s PANSS Positive (p=0.02), PANSS Negative (p=0.045) and PANSS General Psychopathology (p=0.008) scores were significantly lower than controls, as well as higher scores on SAI (schedule for assessment of insight) (43) measuring insight (p=0.01). At 6-month FU, only PANSS Positive scores sustained significant improvement (21).

A pilot RCT for CaCBTp (22) recruited 42 participants with a diagnosis pf schizophrenia. The experimental group (n=12) received 16 sessions of CaCBTp over 4 to 6 months in line with NICE guidelines (44) and aligned with protocol developed by Kingdon and Turkington (42). Adaptations were based evidence from Naeem et al (45). These included involving family from the beginning of treatment, using a bio-psycho-spiritual-social model of mental health, and involving input from faith healers. At the end of therapy, experimental group showed significant improvement on PANSS Positive (p=.000), PANSS Negative, (.000), PANSS general psychopathology (p=.000), PSYRATS (psychotic symptom rating scale) (46) (p=.000) and insight (p=.000) (22).

A RCT for CaCBTp by Naeem et al., (23) recruited 116 participants from 2 hospitals in Karachi, Pakistan. Participants received CaCBTp plus TAU (n=59) or TAU (n=57). The brief CaCBTp encompassed 6 sessions with involvement of main carer and one session dedicated to family over 4 months. Treatment protocol followed the Kingdon and Turkington (42) manual. Preliminary work was conducted via qualitative interviews to establish adaptations. They highlighted barriers in therapy including lack of awareness of therapy; family involvement; distance; expenses and uncooperative caregivers, as well as strengths. The adaptations involved family from the start of therapy, utilised the bio-psycho-spiritual model and sought support from faith and religious healers. The experimental group showed significant improvement on PANSS Positive (p=.000), PANSS Negative, (.000), PANSS general psychopathology (p=.000), PSYRATS hallucinations (p=.000), PSYRATS delusions (p=.000) and SAI (p=.007).

### 3.4 Functional Analytic Psychotherapy (FAP)

#### 3.4.1 India

A study evaluating FAP recruited 10 participants and assigned to either receive FAP (n=5) or TAU (n=5). FAP used techniques such as cognitive rationale, social skills training and applying this in adaptive and community-based environments. Improvements were found in PANSS Positive (p= 0.042), PANSS Negative (p=0.043), general psychopathology (p=0.04) and QOL (p=0.043) and coping skills (CRI-A) (p<.01) (24).

### 3.5 Psychoeducation

#### 3.5.1 India

A single session of psychoeducation experimental study (intervention (n=9) versus control group (n=25)), scored low on implementation readiness (19/52) (25). Experimental group received one hour of psychoeducation covering modules in symptoms, diagnosis, course of illness and prognosis. Issues of stigma, helplessness and social exclusion were covered. Both groups received antipsychotic medication administered by their treating clinicians who were blind to the study. Data was collected at one-month and three-month FU points. There was significant (p<.005) improvement in the experimental group’s PANSS Positive, PANSS total, symptom severity a global assessment of functioning (GAF). No significant change was found in PANSS Negative, SAI and WHOQOL-BREF (quality of life) scores in both groups and no significant change of PANSS general psychopathology in the experimental group. No significant findings were found between groups (25).

Baruah et al., (26) recruited 60 participants with a diagnosis of schizophrenia from the inpatient department of LGB Regional Institute of Mental Health, Tezpur, Assam and assigned to either an experimental (n=30) or control (n=30) group. The experimental group received a psychoeducative intervention package. Participants were observed on performance of self-care and knowledge about the diagnosis. Measures were not standardised, however good reliability (r=.913; r=.70) and validity (five raters used) was found. The experimental group showed significantly higher knowledge scores (p<.05) compared to baseline and controls post-intervention (p<.05). Observed self-care performance was significantly higher (p<.05) compared to controls post-intervention. Post-hoc analysis showed there was no significant relationships between post-test self-care performance and post-test knowledge in the experimental group (p>.05), however due to lack of standardisation and design, it was awarded the lowest IR score (12/52) (26).

### 3.6 Cognitive rehabilitation/ retraining

#### 3.6.1 India

A RCT evaluated the effectiveness of a cognitive retraining programme. All patients and caregivers received psychoeducation and were assessed at baseline, at 2-months and 6-months (27). Forty-five participants with first episode psychosis were randomly assigned to either experimental group receiving home-based cognitive retraining along with TAU (n=22) or TAU only (n=23). The retraining encompassed attention, mental speed, information processing, memory, verbal fluency, planning, visuospatial reasoning. The experimental group showed significant improvement across most cognitive domains (p<.05), PANSS scores (p<.001) and global functioning (WHODAS; p<.001). Significant differences between groups were found for motor speed (p=.019), executive functioning (p=.03), verbal learning and memory (p=.03), visual learning and memory (p=.02) and PANSS negative symptoms (p=.019) (27).

A case study of a 26-year-old female undergoing cognitive retraining following a diagnosis of paranoid schizophrenia was evaluated with a pre-post design over 10 weeks. The intervention consisted of neurofeedback training, cognitive retraining in line with protocol developed by (27) and a family intervention to address interpersonal difficulties, aid the family in supporting the participant and address any issues around caregiver burden. The participant completed retraining tasks at home for 1-hour daily. Significant improvement in social functioning was found, including communication style, motivation, empathy, and emotional reactivity. Significant improvement were found in most cognitive domains excluding abstraction (15).

### 3.7 Social skills training/ social cognition

#### 3.7.1 India

Twenty-nine clinically stable participants with schizophrenia (n=27) or schizoaffective disorder (n=2) according to DSM-IV-TR were recruited from the outpatient department of National Institute of Mental Health and Neurosciences (NIMHANS) and their caregivers. A pre-post design was used to evaluate the effectiveness of an integrated psychological treatment (IPT). IPT involved between 16– 20 bi-weekly sessions over two and a half months. Modules included cognitive differentiation, social perception, verbal communication, and social skills and were modified to the client’s specific needs. Caregivers received psychoeducation about schizophrenia, symptoms, causes adherence, expressed emotion and prognosis which were tailored to the participants’ specific needs. Significant differences were found pre and post for neurocognitive abilities for intelligence quotient (p<.001), colour trails (p=0.027), controlled word association test (p=.004), all domains of Reys auditory verbal learning test (p<.001) and the immediate and delayed recall elements of the Rey’s complex figure test (p<.001). Significant differences were found between pre and post for social functioning (p<.001), PANSS Positive (p<.05), PANSS Negative (p<.001), PANSS general (p<.01) and PANSS total (p<.001) (28)

A culturally adapted 16– 20 session IPT was evaluated over two and half months to 29 clinically stable individuals with a diagnosis of schizophrenia. The intervention was culturally adapted and 23 participants who completed the FU assessments were analysed. Family caregivers received psychoeducation about schizophrenia and was involved in homework tasks. Significant improvement was found in social cognition (p<.001), social skills (p<.001), PANSS Positive (p<.017), negative (p<.001), general psychopathology (p<.007) and total (p<.001). Post-intervention social perception showed a medium effect size (0.77) and large for social skills (0.88). At FU, large effect sizes were found for social perception (0.94), social cue recognition (0.96), social skills (0.92), social and occupational functioning (1.22) (29).

Another study evaluated the effectiveness of the Indian version of the training of affect recognition (TAR) intervention (30), a social cognition training programme with 10 participants with a diagnosis of schizophrenia (International Centre of Disease-10 [ICD-10]) (WHO, 1993) recruited from a rehabilitation centre. The intervention consisted of three blocks of four sessions. Each block covered identifying and verbalising facial features of six basic emotions (happy, sad, fear, anger, surprise, and disgust); identifying and interpreting them in social situations, cues, and attribution bias. All tasks were translated into Kannada, native language of Karnataka state in India, social contexts were rendered to be culturally appropriate and administration methods were modified. There was a significant change in performance on emotion recognition between pre and post intervention (p<.001) measure by Tool for Recognition of Emotions in Neuropsychiatric Disorders (TRENDS). There was no significant change in PANSS symptoms (p>.05) (30).

### 3.8 Caregiver/family interventions

#### 3.8.1 India

Shihabuddeen et al., (31) delivered psychoeducation and problem-solving groups monthly for 17 months. Twenty-three caregivers of individuals with schizophrenia were recruited from a psychiatric rehabilitation unit. Post-intervention, 22 of clients with schizophrenia reported having a good family support system, compared to 4 before the meeting, adequate coping skills compared to 1 prior to the meeting and good treatment compliance post, compared to 0 prior to the intervention. All 23 reported positive attitudes of the caregiver towards the client, compared to 10 before the meeting. Due to lack of standardisation, it scored low on the IR (17/52).

Kumar et al., (32) evaluated a 2-session psychoeducation and group therapy intervention in 66 participants and key relatives of patients over 3-months, followed by bi-weekly follow-up appointments for 3-months. This consisted of rapport building, education about schizophrenia, expressed emotion, role of key relatives, medication support, compliance, coping and problem-solving approaches. They found significant reduction in burden of care (Burden Assessment Scale; BAS) (p=.004), improvement in QOL of caregivers (WHOQOL-BREF) (p=.024) and QOL of patients (p=.0028) in the experimental group. They study scored high on IR (34/52).

Devaramane et al., (33) delivered a brief psychoeducation intervention over 3 sessions and covered education about schizophrenia, communication, problem solving skills and expressed emotion. Twenty patients with schizophrenia (ICD-10 diagnosis) and primary caregivers were recruited from an inpatient and outpatient medical academy in India, 18 completed the study. There was significant improvement in patients PANSS total and positive symptoms (p<.001); PANSS negative and general psychopathology (p<.05); BAS (p<.001) and family crisis oriented personal evaluation scale (F-COPES) (p<.05); it scored moderately on IR (28/52).

Sadath et al., (34) delivered a 7-session psychoeducation intervention included psychosocial management of illness, psychosocial needs of caregivers, engagement, psychoeducation about psychosis to 59 participants with first episode psychosis and their caregivers. Expressed emotion Widermann et al., 2002) in the experimental (p<.001) and TAU (p<.001) groups significantly reduced over time, but no significant difference were found between groups over time (p=.184). Social support improved significantly in both groups over time (experimental p<.001; TAU p<.001) but no significant differences between groups over the period (p=.064).

However, carers in the experimental group showed significant improvement in significant others support (p=.023) and friends support (p=.009) domains of the social support measure. It scored moderately on IR (26/52).

Kulhara et al., (14) evaluated a psychoeducation intervention in 76 participants with a DSM-IV diagnosis of schizophrenia and their caregivers who were randomly assigned to receive either the intervention (n=38) or TAU (n=38) for 9-months. The intervention covered causes of schizophrenia, symptoms, treatment, goal setting, substance abuse, marriage, and communication. Symptoms were monitored monthly, whilst disability, caregiver-burden, caregiver-coping, caregiver-support, and caregiver-satisfaction were assessed pre and post intervention. The intervention group showed significantly more improvement in PANSS scores (p<.001), disability scores (p<.001), caregiver support (p<.001) and satisfaction compared to TAU. No significant differences were found for caregiver burden or coping (p>.05).

Anchan and Janardhana (36) evaluated the effectiveness of psychoeducation on reducing expressed emotion in husbands of women with post-partum psychosis. Three sessions of psychoeducation covered misconceptions about the diagnosis, common symptoms, biopsychosocial model explanations and their role in managing the condition whilst supporting themselves. They attended on a daily or weekly basis; assessments were conducted immediately after completion and at 1-month FU. Significant improvements were found in authoritarianism when comparing pre to post intervention (p<.05), and pre-intervention to FU (p<.05); pre- and post-intervention social restrictiveness (p<.05) and pre to FU (p<.05); pre- and post-intervention community mental health ideology (p<.05) and pre to FU (p<.05).

Chakraborty et al (37) evaluated an 8-session psychoeducation session for caregivers. This comprised of course, prognosis, outcomes, emotional and behavioural effects on patients, treatment, side effects, stressors, the role of the family in rehabilitation and problem solving. Fifty caregivers with one family member with a diagnosis of schizophrenia (simplex) and 30 with multiple (multiplex). They found significant reductions in the simplex group across all domains of the BAS (p<.001), effect sizes (−0.6- −3.8), and the problem solving, distraction, acceptance, and denial domains of coping skills (p<.001), effect sizes (−2.6-2.81). In the multiplex group, the spouse related, physical and mental health, caregiver routine, support of patients, taking responsibility, patients’ behaviour, caregiver strategy were all significant (p<.001) effect sizes (−4.8- −1.6), and problem solving, distraction, acceptance, and denial domains of coping skills (p<.001), effect sizes (−4.8- −2.3).

#### 3.8.2 Pakistan

Nasr et al., (35) evaluated a psychoeducation package for 108 patients with a diagnosis of schizophrenia and their caregivers recruited from an outpatient department. The intervention followed protocol set out by Kuipers et al., (47) and encompassed information about schizophrenia, causes, treatment and the family’s role in recovery. Both groups received antipsychotics, one group received psychoeducation with 99 completing the study. Burden Interview Schedule (48) scores showed significant reduction in burden in the experimental group compared to controls in financial (p=.056), routine (p=.01), leisure (p<.001), interaction (p<.001) and psychological health (p=.035).

### 3.9 Telehealth intervention

#### 3.9.1 Afghanistan

A cross-sectional study was conducted to assess outcomes of group psychoeducational meeting interventions across 7 districts and 3 control districts. One hundred and fifty meetings were conducted in intervention districts attended by >10,000 people. Significantly more people in the intervention group denied myth of ‘jins’ (evil spirits), jadu (black magic) or “Wrath or God” cause psychosis compared to controls (p<.001). Significantly more intervention participants found the text messages easy to understand (p<.001) and should be continued (p<.001) at point of evaluation compared to baseline (38).

### 3.10 Yoga-based intervention

#### 3.10.1 India

Varambally et al (39) recruited 29 caregivers of people with psychosis (ICD-10) and randomly assigned them to yoga (n=15) or waitlist (n=14) conditions. Assessments were taken pre-intervention and 3-month FU, the intervention group received yoga training thrice week for 4 weeks and instructed to practice for the following 2-months. BAS scores were significantly reduced (p<.05) and WHOQOL-BREF scores improved significantly (p<.05) at FU compared to controls. No significant changes were found in psychopathology and disability in patients or anxiety and depression scores in caregivers.

## 4. Discussion

To date, to the best of our knowledge, no other systematic review addresses the effectiveness and implementation readiness of a range of psychosocial interventions or programmes for supporting people with psychosis in South Asia. Twenty-six articles evaluating nine different interventions across five countries in South Asia were identified. They were situated in India, Pakistan, Afghanistan, Nepal and Sri Lanka. Populations consisted of those with a diagnosis of schizophrenia or any variant of psychosis with psychotic features diagnosed using either the ICD-10 or DSM-IV-TR criteria, or PANSS. The quality of included studies was variable, as were the interventions and outcomes used. Most notably, the highest score awarded for implementation readiness was 42/52, with almost all studies failing to consider or report levels of managerial or employee support for the intervention and few studies reported training costs, time needed for planning and setting up the intervention or barriers to implementation. Commitment, involvement and accountability of managers is an essential component of a conducive implementation climate and increases the likelihood of successful implementation of an intervention (49).

Significant improvement in psychological factors (e.g., symptoms) was found in eight studies. These were multi-component community-based interventions in India, CaCBTp interventions in Pakistan, family interventions in India and a cognitive retraining programme in India.

MCBI encompassing psychoeducation, caregiver support and treatment adherence yielded similar results (16). This study encompassed structured needs assessments, psychoeducation, adherence management strategies, individualised rehabilitation strategies to improve the person, social and occupational functioning. It addresses stigma and discrimination issues, social inclusion, access to benefit and employment schemes and involves caregivers throughout. Moreover, community health workers were able to deliver the intervention. This provides promising evidence that interventions addressing multiple components including social and life-skills yield the most effective results in the treatment of psychosis across countries in South Asia. Although they initially increase in costs, research suggests that interventions that address symptoms, involve the wider social network and promote social and occupational functioning are effective at preventing relapse, thus longitudinally can reduce costs in financial burden of the family and healthcare system. Furthermore, the outcomes are consistent with evidence from Gopal et al. (50) who found social and life-skills to be integral parts of recovery for those with psychosis in LMICs and should be integrated in treatment packages.

Other studies providing community-based interventions/ assertive outreach found significant reductions in disability and symptoms, caregiver burden obtained higher ImpRess checklist ratings and were studied in different countries including India, Nepal and Sri Lanka (18, 19, 20). These studies also utilised mental health development officers and primary care mental health workers further strengthening the argument for a task shifting model and utilising the existing workforce. Supervised community health workers have been advocated for as a way to scale up services when specialist resources are scarce in LMICs (51). They can provide accessible, community-based services to people with psychosis. Although Chatterjee et al., (16) reported having to complete 6-weeks training, this was due to the RCT guidelines, and suggest this could be cut down to 3-weeks in a real-world setting. The findings suggest the model is most effective for those with moderate to severe schizophrenia, by task-shifting to lay workers, specialist resources are then more accessible for those who could be more difficult to treat, in line with standard practice in high-income countries and WHO and the Expert Policy Group of the Ministry of health in India.

CaCBTp scored well on the ImpRess checklist. The cultural adaptations increase the likelihood that the intervention is implemented into standard practice. However, the evidence was limited to Pakistan, so was not deemed ready to be implemented widely in other South Asian countries. One of the key adaptations was reference to religious teachings and religion varies widely within South Asia and might influence findings as well as cultural adaptations.

### 4.1 Limitations

Firstly, the broad inclusion criteria which led to the inclusion of heterogeneous interventions and outcomes meant that a meta-analysis could not be conducted. The Downs and Black items required a fair amount of interpretation from raters. Few studies included in this adopted a randomised-controlled design, however this is not always the most appropriate design for psychotherapy and due to the small sample sizes of some of the studies, randomization would not be appropriate (52).

The ImpRess checklist was originally developed for cognitive stimulation therapy in the UK which questions its validity in assessing interventions for psychosis.

However, other studies have found it to be a useful means of measuring the IR of psychosocial interventions in LMICs (53) and is considered a feasible means of comparing the implementation readiness of psychosocial interventions in general and in different healthcare systems.

Most measures used in included studies were standard for psychosis research. Commonly, studies used PANSS, QOL, WHODAS and BAS. Few studies used culturally adapted measures such as IDEAS. However, three studies used non-standardised measures to assess some or all the outcomes with only one reporting good reliability and validity.

### 4.2 Future recommendations

The findings suggest the implementation of MCBI demonstrates utilisation of limited resources, to upskill lay health workers, can be considered cost-effective long-term and aims to address the needs of the population. The broad inclusion criteria and the fact that participants with a need for additional community care were recruited from real-world clinical settings across three diverse sites and practice arrangements in India all suggest that the findings of the trial are generalisable to similar settings in other low-income and middle-income countries. A review of MCBI, compared across clinical setting-community vs facility based would establish what specific areas and using culturally adapted outcome measures to ensure research is measuring what is appropriate and important in that culture. CaCBTp scored high on implementation readiness due to cultural adaptations within Pakistan. Future research should aim to examine its applicability in other South Asian regions.

Very few studies included in this review reported on the implementation aspects of introducing interventions across countries and cultural contexts. Future research on the effectiveness of psychosocial interventions in South Asia and the LMIC context would benefit from the routine inclusion of implementation readiness data. This will inform clinical practice and policy, aiding the scaling up of interventions to widespread implementation.

### 4.3 Conclusion

MCBI was found to be an adequate intervention for people with psychosis in some countries in South Asia as demonstrated by its efficacy and implementation readiness. The findings suggest improvements in symptomatology, disability functioning and given its utilisation of limited resources and upskilling of lay health workers could address the treatment gap in South Asia in a cost-efficient manner. CaCBTp and a community-outreach intervention were also deemed effective but not ready to be implemented in multiple settings.

## Data Availability

All relevant data are within the manuscript and its Supporting Information files.

## Notes

### Competing Interest Statement

The authors have declared no competing interest.

### Funding Statement

The author(s) received no specific funding for this work.

